# Rare genetic variants in *SEC24D* modify orofacial cleft phenotypes

**DOI:** 10.1101/2023.03.24.23287714

**Authors:** Sarah W. Curtis, Jenna C. Carlson, Terri H. Beaty, Jeffrey C. Murray, Seth M. Weinberg, Mary L. Marazita, Justin L. Cotney, David J. Cutler, Michael P. Epstein, Elizabeth J. Leslie

**Affiliations:** Department of Human Genetics, Emory University, Atlanta, GA, 30322, USA; Department of Human Genetics, University of Pittsburgh, Pittsburgh, PA, 15621, USA; Department of Biostatistics, University of Pittsburgh, Pittsburgh, PA, 15261, USA; Department of Epidemiology, Johns Hopkins Bloomberg School of Public Health, Baltimore, MD, 21205,USA; Department of Pediatrics, University of Iowa, Iowa City, IA, 52242, USA; Center for Craniofacial and Dental Genetics, Department of Oral and Craniofacial Sciences, University of Pittsburgh, Pittsburgh, PA, 15261, USA; Department of Genetics and Genome Sciences, University of Connecticut, CT, 06030, USA

## Abstract

As one of the most common structural birth defects, orofacial clefts (OFCs) have been studied for decades, and recent studies have demonstrated that there are genetic differences between the different phenotypic presentations of OFCs. However, the contribution of rare genetic variation genome-wide to different subtypes of OFCs has been understudied, with most studies focusing on common genetic variation or rare variation within targeted regions of the genome. Therefore, we used whole-genome sequencing data from the Gabriella Miller Kids First Pediatric Research Program to conduct a gene-based burden analysis to test for genetic modifiers of cleft lip (CL) vs cleft lip and palate (CLP). We found that there was a significantly increased burden of rare variants in *SEC24D* in CL cases compared to CLP cases (p=6.86×10^−7^). Of the 15 variants within *SEC24D*, 53.3% were synonymous, but overlapped a known craniofacial enhancer. We then tested whether these variants could alter predicted transcription factor binding sites (TFBS), and found that the rare alleles destroyed binding sites for 9 transcription factors (TFs), including Pax1 (p=0.0009), and created binding sites for 23 TFs, including Pax6 (p=6.12×10^−5^) and Pax9 (p= 0.0001), which are known to be involved in normal craniofacial development, suggesting a potential mechanism by which these synonymous variants could have a functional impact. Overall, this study demonstrates that rare genetic variation contributes to the phenotypic heterogeneity of OFCs and suggests that regulatory variation may also contribute and warrant further investigation in future studies of genetic variants controlling risk to OFC.

## Introduction

Orofacial clefts (OFCs) are one of the most common structural birth defects, affecting 1 in 700 births (Ciminello et al. 2009; Dixon et al. 2011; Gundlach and Maus 2006; Mossey et al. 2009; Mossey and Modell 2012; Sabbagh et al. 2012). OFCs require multiple surgeries early in life, and affected individuals can experience speech, hearing, and psychosocial problems throughout life (Hameed et al. 2019; Lancaster et al. 2020; Wehby and Cassell 2010; Wehby et al. 2006). A majority (∼50-70%) of OFCs cases are classified as nonsyndromic and most of these nonsyndromic cases are hypothesized to be caused by complex or multifactorial etiologies (Leslie and Marazita 2013), having both environmental (Bell et al. 2014; Candotto et al. 2019; DeRoo et al. 2016; Jamilian et al. 2017; Li et al. 2010; Martelli et al. 2015; Yin et al. 2019) and genetic risk factors (Birnbaum et al. 2009a; Birnbaum et al. 2009b; Bishop et al. 2020; Duan et al. 2020; Hao et al. 2018; Leslie et al. 2017a; Leslie et al. 2017b; Leslie et al. 2016; Leslie and Marazita 2013; Li et al. 2020; Li et al. 2017; Ludwig et al. 2016; Ludwig et al. 2017; Mangold et al. 2010; Mukhopadhyay et al. 2020; Shaffer et al. 2019).

OFCs are also phenotypically heterogeneous and can be classified as a cleft of the lip (CL), cleft of the palate (CP), or a cleft of both the lip and the palate (CLP). A majority of the previous epidemiological and genetic studies have grouped types of OFCs together for increased statistical power under a hypothesis of shared biology (Ciminello et al. 2009; Dixon et al. 2011; Leslie et al. 2016; Leslie and Marazita 2013). CL and CLP are most commonly grouped together because of a shared 2:1 male-to-female sex bias and the fact that formation of the lip precedes the formation of the palate during embryonic development (Amidei et al. 1994; Marazita 2012; Urbanova et al. 2013). However, while genetic studies have successfully identified loci that contribute to both CL and CLP (Beaty et al. 2010; Birnbaum et al. 2009a; Birnbaum et al. 2009b; Leslie et al. 2016; Mukhopadhyay et al. 2020), recent studies indicate that CL and CLP can also have distinct genetic risk factors (Carlson et al. 2019; Carlson et al. 2017a; Carlson et al. 2017b; Curtis et al. 2021a; Curtis et al. 2021b; Huang et al. 2019), or genetic modifiers, that predispose for the formation of one cleft type over another. We previously performed a targeted sequencing study of known OFC-associated regions and found evidence that common variants near 9q22, 17p22, and 20q12 are associated with the formation of CL or CLP and rare variants in 7 genes are associated with cleft type (Carlson et al. 2017b). Additional targeted studies have shown that some genotyped SNPs in *IRF6* have a stronger effect in CL compared to CLP (Carlson et al. 2019). Genome-wide, subtype specific effects have been found for CL (Huang et al. 2019), and we also showed in a genome-wide modifier analysis that common variants in 16q21 are more strongly associated with the formation of a CL over a CLP, as are low-frequency variants in *C8orf34, TMEM246*, and *CDC42EP3* (Carlson et al. 2017a).

These studies indicate that common and low-frequency genetic variants are associated with the phenotypic variability among OFCs. However less is known about the contribution of rare variants genome-wide to the phenotypic heterogeneity of OFCs given the relatively limited scope of previous studies of rare variants (Carlson et al. 2017b). Therefore, we set out to test whether any genes had a burden of rare variants using whole genome sequencing data from the Gabriella Kids First Pediatric Research Program, focusing on CL vs CLP cases.

## Methods

### Cohort Description

This study used affected probands with cleft lip (CL) or cleft lip and palate (CLP) and their parents that were sequenced as part of the Gabriella Miller Kids First Pediatric Research Program (GMKF). Participants were recruited from the United States, Argentina, Turkey, Hungary, Spain, Colombia, and Taiwan. Participant recruitment was done at regional treatment or research centers after review and approval by each site’s institutional review board (IRB) and the IRB of the affiliated US institutions (e.g., the University of Iowa, the University of Pittsburgh, and Johns Hopkins University). Because we were interested in genetic differences in CL vs CLP cases and 93.5% of the CL cases were in the samples with European ancestry, only the subset of samples with European ancestry were retained for analysis. For subtype-specific analyses (described below), 76 trios with isolated CL and 225 with CLP were used. The 20 trios (representing 6.6% of the dataset) with affected parents were retained in this analysis, but trios from families with both CL and CLP were excluded. For the modifier analysis (described below), we randomly selected a subset of unrelated individuals, with 100 having CL and 269 having CLP. For both analyses, the proportion of male and female probands were similar: 67% male, 33% female for the subtype-specific analysis and 66%, 34% female for the modifier analysis (Table S1).

### Variant calling and QC

The whole-genome sequencing and quality control methodology for these cohorts has been described in detail previously (Bishop et al. 2020; Mukhopadhyay et al. 2020). Briefly, DNA from either blood or saliva samples were sequenced (depending on availability) at either the McDonnell Genome Institute (MGI), Washington University School of Medicine in St. Louis or the Broad Institute. Variants were called and aligned to the GRCh38/hg38 reference genome using the GATK pipeline (Conrad et al. 2011; McKenna et al. 2010; Van der Auwera et al. 2013) at the GMKF Data Resource Center. The details of how these datasets were aligned and genotyped and harmonized has been previously published (Bishop et al. 2020; Mukhopadhyay et al. 2020). For sample-level QC, individuals with missingness or Mendelian error rare, or average read depth outside of three standard deviations from the mean were removed, as were individuals with a transition/transversion ratio (Ts/Tv), exonic Ts/Tv, silent/replacement ratio, or heterozygosity/homozygosity ratio outside expectation (Bishop et al. 2020). PLINK (v1.9) (Chang et al. 2015) was used to calculate identity-by-descent (IBD) to confirm all family relationships and X chromosome heterozygosity to confirm the sex of all participants. For variant-level QC, non-passing variants, variants with more than 2 Mendelian errors, variants with >5% missingness in individuals, variants with a Hardy-Weinberg equilibrium p-value < 1×10^−7^ in unaffected parents, and variants with a quality by depth (QD) score of <4 were removed. Additionally, calls with a genotype quality (GQ)<20 or a depth (DP)<10 were set to missing and only biallelic variants were retained using VCFTools (Danecek et al. 2011).

### Annotation and filtering

All variants were annotated to publicly available databases using ANNOVAR (v201707) (Wang et al. 2010) and Variant Effect Predictor (VEP) (McLaren et al. 2016). Variants were defined as rare if their minor allele frequency (MAF) was less than 1% or 0.1% in all populations in gnomAD (v3.1.2) (Karczewski et al. 2020), and the allele count (AC) in this population was <42 (for MAF 1%) and <8 (for MAF 0.1%). Any coding (protein-altering or synonymous) variant was retained for analysis.

### Statistical analyses

Subtype-specific analyses were performed by testing for a burden of rare variants genome-wide using a rare variant extension of the transmission disequilibrium test (RV-TDT) (He et al. 2014) in trios with just CL (N = 76) and trios with just CLP (N = 225) separately. Variants were collapsed by RefSeq-defined gene region, and each gene with 2 or more variants was tested. Any variant that was coding and had a MAF < 1% in gnomAD (v3.1.2) was included in the analysis. We used a Bonferroni threshold correcting for the number of genes tested to determine statistical significance (CL analysis: p < 3.83×10^−6^; CLP analysis: p < 2.86×10^−6^). The - log_10_ of the p-values from the two analyses were compared with a Pearson’s correlation.

A modifier analysis for CL vs CLP was performed using RVTESTS (Zhan et al. 2016). This approach has higher power to identify genetic risk factors that differ between two subtypes, but no power to find factors important in both groups. We directly compared an unrelated subset of CL (N = 100) and CLP (N = 269) cases. Rare variants were collapsed across genes, and the association was tested using the Combined and Multivariate Collapsing method (CMC), which calculates the cumulative burden of rare variants within a region and is designed to find sets of rare variants with the same direction of effect. Any variant that was coding and had a MAF < 1% in gnomAD (v3.1.2) was included in the analysis. A Bonferroni threshold to correct for the number of genes tested was used to determine statistical significance (0.05/19,208 genes; p < 2.60×10^−6^). Additionally, we considered p-value < 1×10^−5^ to be suggestive and performed additional analyses to localize the signal. These analyses included restricting variants to those that were rarer (MAF <0.1%), only protein-altering, or including coding and non-coding variants in specific regions (enhancer region definitions defined below). Sex and the first 8 principal components (PCs) were used as covariates. PCs were calculated using the GENESIS software package (Gogarten et al. 2019).

### Functional annotation of results

Topologically-associated domains (TADs) were defined for significantly associated loci using the H1-ESC cell line in 3D Genome Browser (Wang et al. 2018). Regions with potential craniofacial enhancers were annotated using publicly available datasets derived from ChIP-seq data from human embryos at CS13, CS14, CS15, CS17, and CS20 (4.5-8 weeks post conception) (Attanasio et al. 2013; Wilderman et al. 2018), and enhancers that have been experimentally validated as part of the VISTA Enhancer Browser (Visel et al. 2007). Gene expression for the significant and suggestive genes from the analyses was assessed with RNA-seq data generated from human cranial neural crest cells (GEO: GSM1817212, GSM1817213, GSM1817214, GSM1817215, GSM1817216, and GSM1817217) (Edgar et al. 2002; Prescott et al. 2015), and throughout craniofacial development (CS13-CS20) (Yankee et al. 2022). For any variants within a predicted enhancer region that was suggestive in the main analysis, transcription factor binding sites (TFBS) were predicted using TFBSTools (Tan and Lenhard 2016) and the JASPAR database (Khan et al. 2018). The 50 base pairs surrounding each variant within the region were extracted from the reference sequence, and the analysis was conducted using both the reference sequence and the alternate sequence. We then selected predicted TFBS that both overlapped with the position of the variant and differed between the reference and alternate sequence. TBFS were considered predicted if the p-value was significant after multiple test correction (p < 0.05/Number of TFBS that overlapped the variant position). The TFBS were considered different between reference and alternate sequence if the TFBS was significant in one sequence and not the other.

## Results

We performed a subtype-specific genome-wide analysis to identify genes with a burden of rare variants by performing a RV-TDT analysis in trios with just cleft lip (CL) and trios with just cleft lip and palate (CLP), separately. This type of analysis could detect variants associated with increased risk for an OFC in general, but could also identify variants that increase the risk for just one of the subtypes of OFCs. Overall, no gene was significant in either analysis (Figure S1-S2, Table S2-S3), likely due to the small sample size. However, some of the top results were in genes known to be associated with OFCs in previous analyses of common variants (e.g., *ARHGAP29*: p = 0.002 in the CL analysis)(Beaty et al. 2010; Leslie et al. 2016; Yu et al. 2017). The top results for CL and CLP were different and the p-values were not correlated (r = 0.005, p = 0.56; Figure S3). Only one gene had a p-value < 0.01 in both the CL and CLP analysis (*CYP2D7*: p = 0.007 in CL, p = 0.005 in CLP), with 52 genes being p < 0.01 in CL alone and 108 genes being p < 0.01 in CLP alone. This result is consistent with the hypothesis that CL and CLP may have partially distinct genetic architectures.

Because of the differences between the burden results from CL and CLP, we next performed a modifier analysis by comparing CL cases directly to CLP cases. Because this is a case-to-case group comparison, this analysis would not be able to detect variants generally important for OFC risk, but would detect variants important for the formation of one subtype over the other subtype. In the modifier analysis, *SEC24D* was genome-wide significant (p = 6.86 10^−7^), and *SUPT4H1* was suggestive (p = 6.33 10^−6^, Figure 1, Figure S4, Table 1, Table S4), with both having a burden of rare variants in CL compared to CLP (Table S5-S6). Both genes were still significant if only rarer variants (MAF < 0.1%) were included in the analysis (*SEC24D*: p = 7.58 10^−5^; *SUPT4H1*: p = 0.001) and if only protein-altering variants were included (*SEC24D*: p = 0.003; *SUPT4H1*: p = 0.0008), indicating the result is robust to different filtering and variant classification strategies.

**Figure 1:**
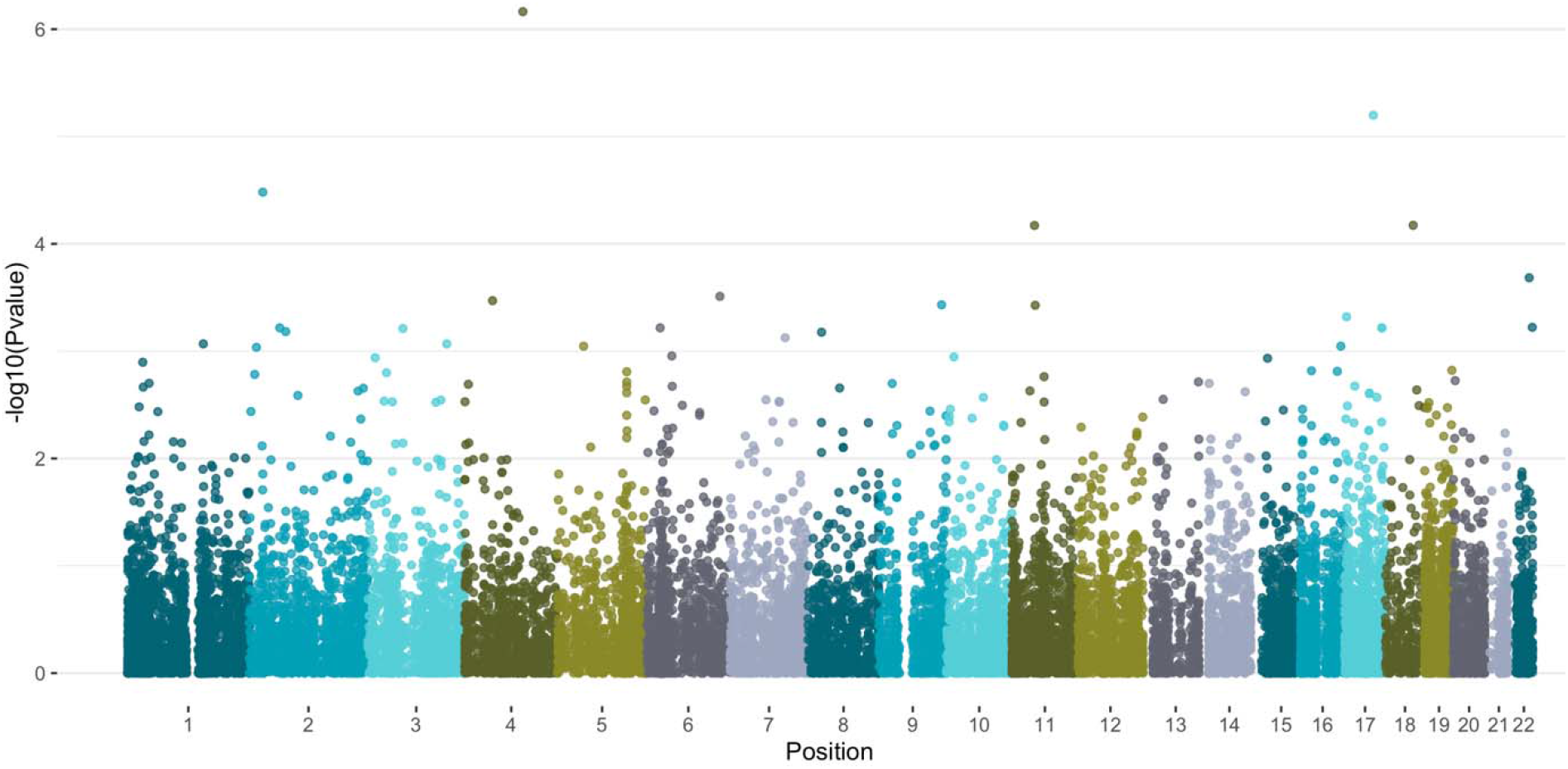
Manhattan plot of –log_10_(p-values) from the cleft lip vs. cleft lip and palate modifier analysis.

**Table 1.** Top modifying genes for CL vs. CLP.

| Table 1. Top modifying genes for CL vs. CLP |  |  |  |  |  |
| --- | --- | --- | --- | --- | --- |
| Gene | Chromosome | Start (hg38) | End (hg38) | Number of rare variants | P-value |
| *SEC24D* | 4 | 118722823 | 118838683 | 16 | $6.86 \times 10^{-7}$ |
| *SUPT4H1* | 17 | 58345175 | 58353093 | 2 | $6.33 \times 10^{-6}$ |
| *C2orf16* | 2 | 27537386 | 27582721 | 49 | $3.30 \times 10^{-5}$ |
| *TXNL1* | 18 | 56597209 | 56651600 | 5 | $6.72 \times 10^{-5}$ |
| *C11orf94* | 11 | 45906513 | 45907272 | 9 | $6.74 \times 10^{-5}$ |

We next further investigated these genes to better understand their role in craniofacial development by analyzing their expression during relevant timepoints, where the rare variants fell within the genes (Figure 2A, Figure S5), and the predicted topological domain (TAD) for both regions (Figure 2B, Figure S6). *SEC24D* and *SUPT4H1* were highly expressed in an *in vitro* human cranial neural crest cell model and throughout human fetal development, being in the top 10^th^ percentile of genes expressed in cranial neural crest cell lines and in the 20^th^ percentile of all genes expressed throughout development (Table S7), further indicating that these are both genes with a potential role for the formation of an OFC. Rare variants were distributed throughout *SEC24D*, including overlapping with a predicted craniofacial enhancer (Figure 2). Rare variants in *SUPT4H1*, however, were just in the beginning of the gene, and within a craniofacial enhancer that covered the entire gene (Supplemental Figure).

**Figure 2:**
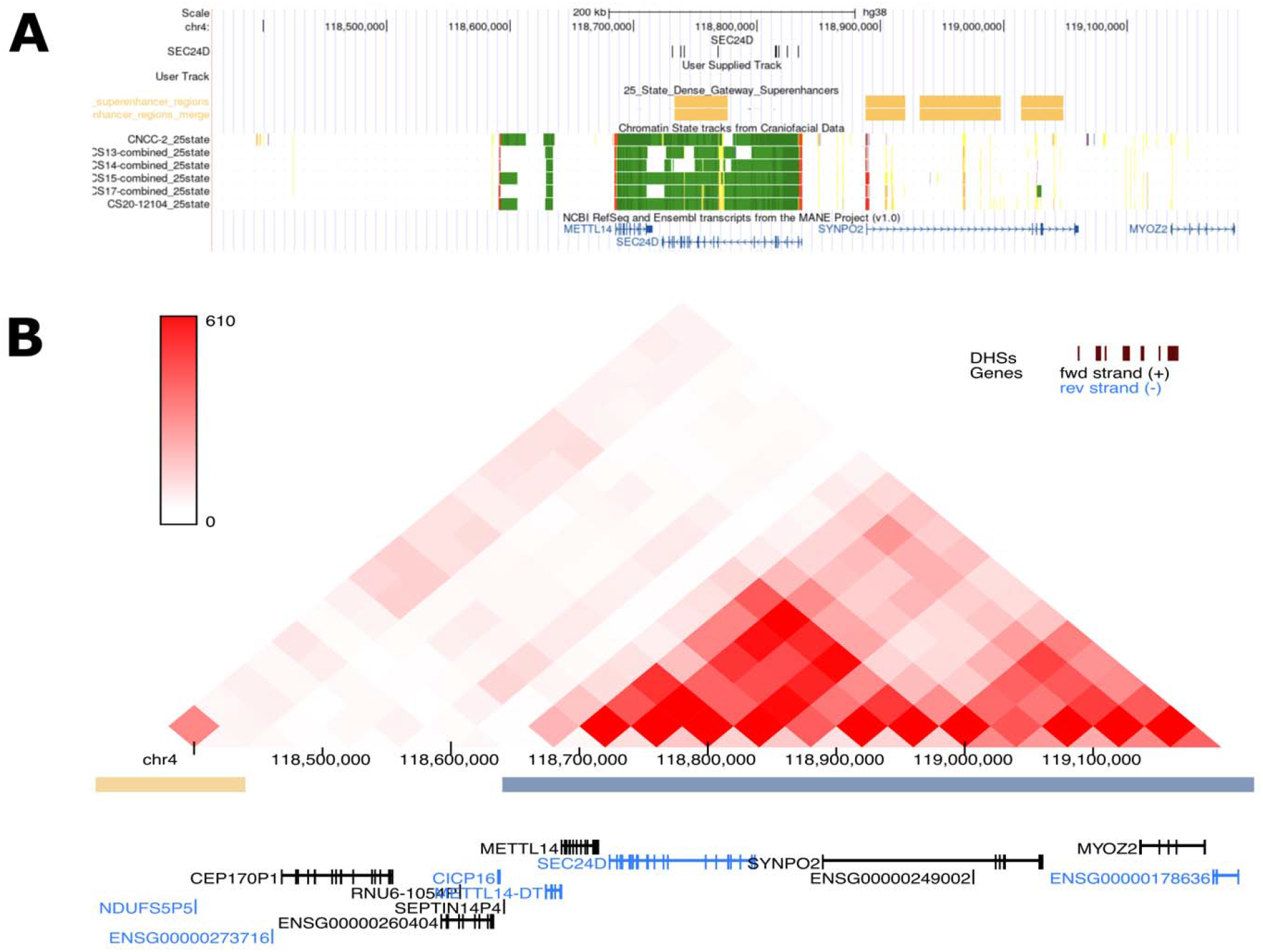
Rare variants within *SEC24D* fall within a predicted craniofacial super enhancer. (A). The predicted topologically-associated domain (TAD) surrounding SEC24D (B).

Because some of the variants overlapped predicted craniofacial enhancers, we next investigated whether the burden test was actually detecting a non-coding signal. We tested the enhancers within the *SEC24D* region to see if there was a burden of either coding (Table S9) or any (Table S10) rare variants. A super enhancer had a burden of rare variants when considering both coding variants (p=0.02) and all variants (p=0.006), indicating that there may be a statistically significant role for non-coding variants, but to a lesser degree than coding variants in this region. We next tested whether any of the variants affected predicted transcription factor binding sites (TFBSs). For *SEC24D*, 9 binding sites for 9 transcription factors (TFs) were destroyed, including ones for PAX1, and 23 TFBS were created, including ones for Pax6 and PAX9 (Table 2, Table S8), suggesting a potential mechanism for a non-coding effect. Within the predicted TAD that contains *SEC24D, SEC24D* was the highest expressed gene, and thus the best candidate target gene for these enhancers, with the next highest expressed gene being *SYNPO2* (Table S7).

**Table 2:**
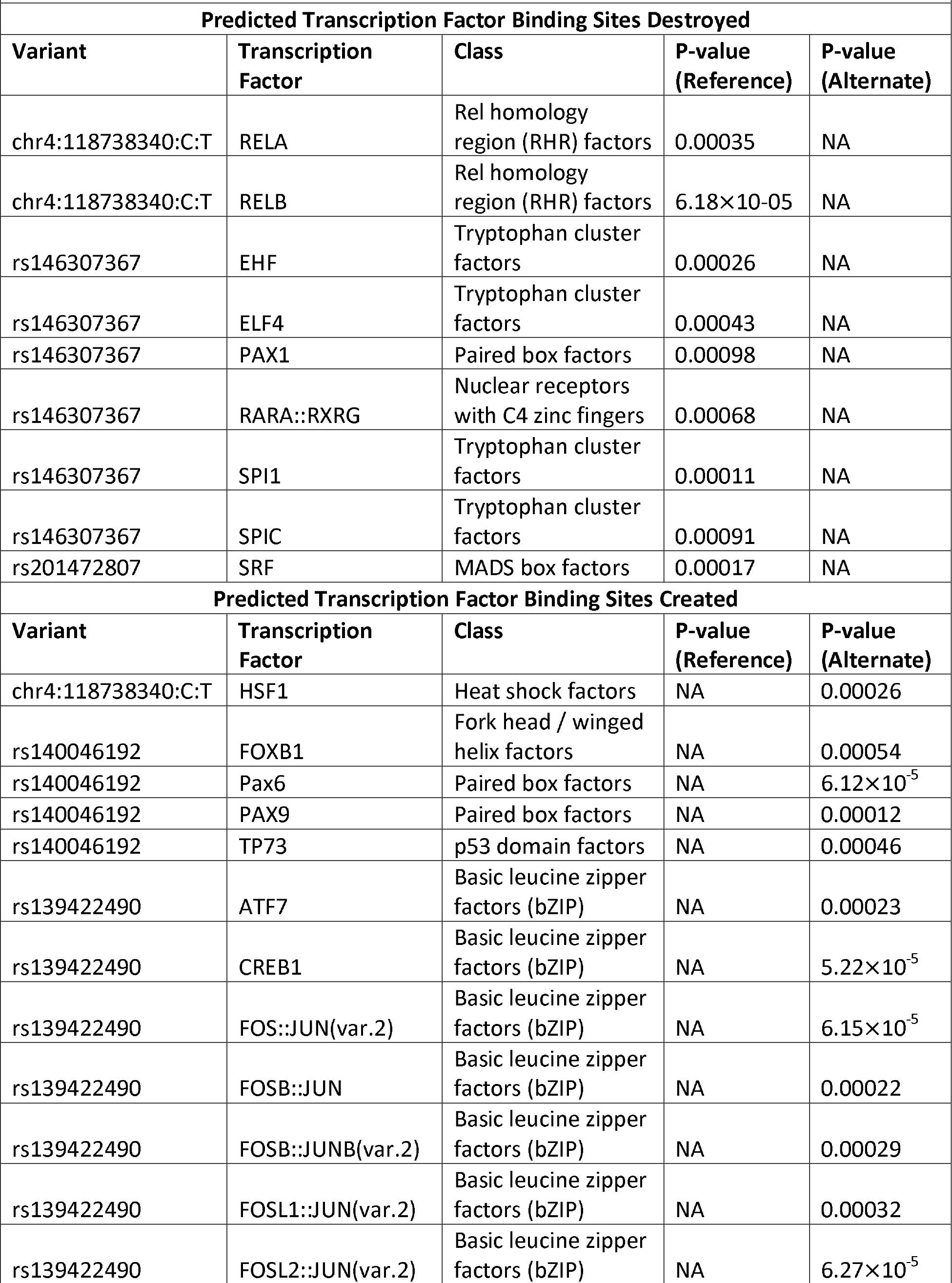
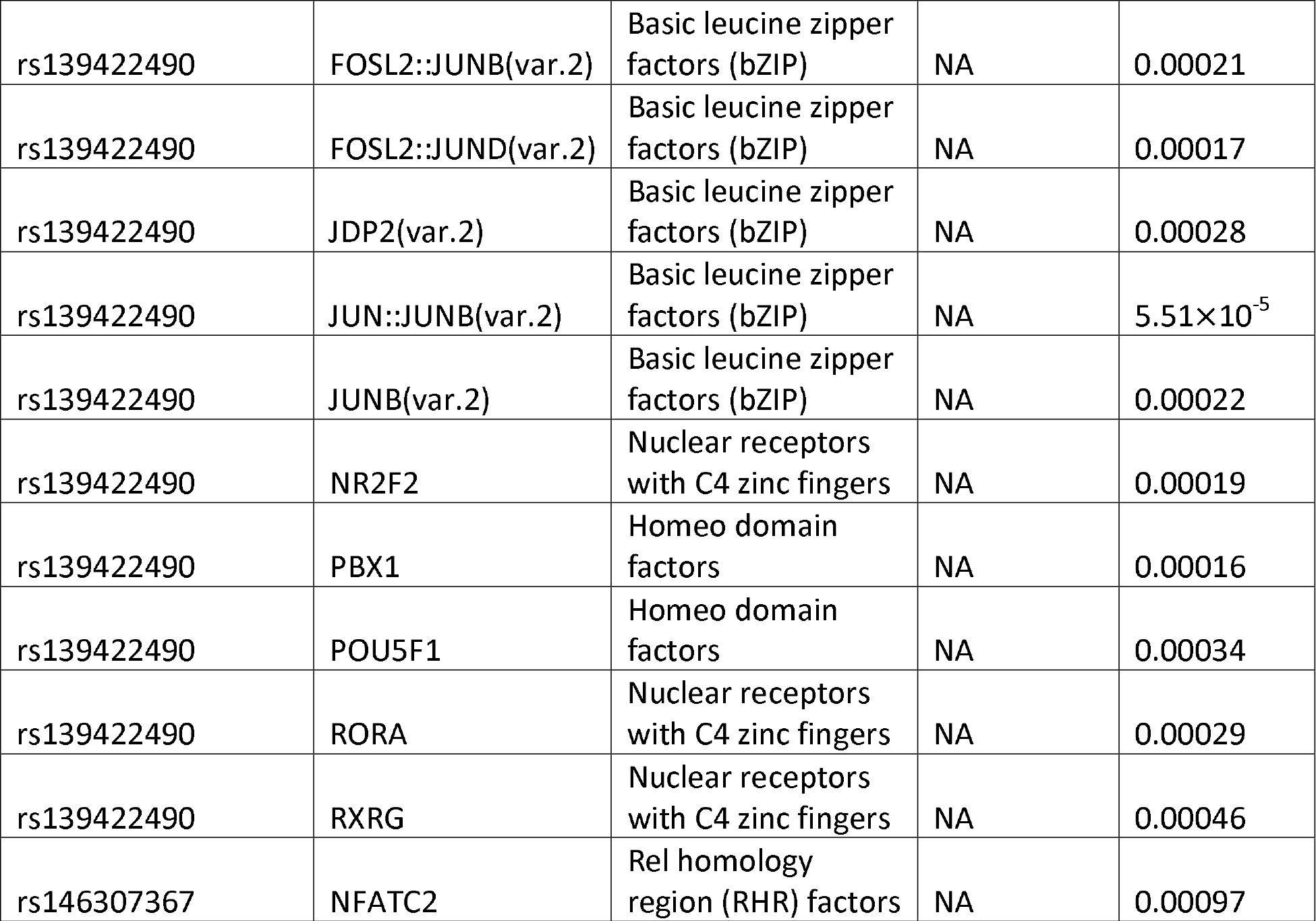
Transcription factor binding sites altered by SEC24D variants within enhancer regions

For the enhancers that overlapped *SUPT4H1*, the super enhancer was significant with just the coding variants (p=6.33 × 10^−6^), but not when all variants were considered (p = 0.14). Two variants were predicted to create binding sites for 5 TFs and destroy the 9 TFBSs, including FOXP2, which is involved in craniofacial morphology and speech (Table S11) (Xu et al. 2018). Of the other genes in the predicted TAD containing this super enhancer, *MTMR4* and *TSPOAP1* were also highly expressed throughout development (Table S7). Taken together, while it is possible that these rare variants could have non-coding effects, the association with the individual enhancers and expression of the nearby genes are not strongly associated with cleft subtypes.

## Discussion

There have been many studies that have investigated the genetic architecture of OFCs but our understanding of the factors that contribute to formation of CL versus CLP is incomplete, especially for rare variants. We previously addressed this by performing modifier analyses using targeted sequencing data and in low frequency variants from genotyping array (Carlson et al. 2017a; Carlson et al. 2017b), but this is the first study to investigate rare genetic modifiers of CL and CLP genome-wide using whole genome sequence data. We found that there was a statistically significant burden of rare, coding variants in *SEC24D* in participants with CL compared to CLP, and that this association still held if we lowered the minor allele threshold or only included protein-altering variants. We also found from publicly available RNA-seq data that *SEC24D* is highly expressed in neural crest cells lines and during craniofacial development. *SEC24D* is involved in vesicle trafficking, as a member of Coat Protein Complex II (COPII) and is important for cargo selection, concentration, and shape of the vesicle (D’Arcangelo et al. 2013). In zebrafish, *sec24d* is required for the effective transportation of proteins to the extracellular matrix in cartilage and with embryos deficient for *sec24d* have a failure of chondroblast differentiation leading to severe craniofacial dysmorphology and malformed cartilage (Sarmah et al. 2010). Consistent with that, *Sec24*-null mice have early embryonic lethality (Baines et al. 2013). *SEC24D* has also been implicated in bone morphology disorders in humans before, with homozygous or compound heterozygous truncating and missense mutations causing Cole-Carpenter Syndrome (MIM# 616294), a syndromic form of osteogenesis imperfecta that presents with disturbed ossification of the skull, leading to midface hypoplasia, macrocephaly, and micrognathia, as well as brittle bones that break easily (Garbes et al. 2015; Moosa et al. 2016; Takeyari et al. 2018; Zhang et al. 2017). Collectively, this is strong evidence that SEC24D is involved in bone and cartilage morphology, especially during craniofacial development. Therefore, heterozygous coding variants that subtly alter this gene’s function or non-coding variants that alter its expression could modify craniofacial phenotypes, such as whether a cleft of just the lip forms compared to a cleft that also affect the bony palate. However, more work is warranted to investigate how this gene could specifically modify orofacial cleft phenotypes.

Even though it did not meet the significance threshold for this study, we also had suggestive evidence for *SUPT4H1* as a modifier of CL vs CLP. *SUPT4H1* is involved in mRNA processing and transcription elongation, and depletion of *SUPT4H1* in cell lines leads to global reduction in RNA levels; mice lacking Supt4h1 do not survive to birth (Cheng et al. 2015; Naguib et al. 2019). Although not previously associated with orofacial phenotypes, it is highly expressed during craniofacial development and overlaps regulatory regions for craniofacial tissue. Therefore, future studies either trying to replicate this result or investigating the role of *SUPT4H1* in facial development or bone morphology are of interest.

We also conducted a subtype specific RV-TDT, comparing the transmission of rare variants in CL and CLP separately. Nothing in that analysis approached genome-wide significance, likely due to the small sample size and that the subtype-specific analysis would have less statistical power to detect loci that differ between two subtypes than the modifier analysis. However, some of the top results in the subtype specific analyses, like *ARHGAP29*, had been previously associated with OFCs (Leslie et al. 2016), and as cohorts increase in size, we expect to identify more robust signals that will facilitate more precise subtype comparisons.

We previously found a burden of low frequency genotyped SNPs in *C8orf34, TMEM24*6, and *CDC42EP3* (Carlson et al. 2017a), and burden of rare variants around the loci containing *PAX7, ARHGAP2*9, 8q24, *FOXE1, VAX1, NTN1*, and *NOG* (Carlson et al. 2017b). None of these genes/regions were significant or suggestive in this analysis, but this could be due to several factors. For example, previous studies used much larger sample sizes. A larger sample size would be better powered to detect rare variants, and therefore there may be genetic modifiers that were previously identified that this study is not powered to detect. Additionally, this lack of replication also be due to heterogeneous effects of rare variants that are undetectable when using the CMC method. There are also ancestry differences between the studies, with this study only having samples with European ancestry and previous studies being predominantly non-European; any modifier that differs by ancestry may not be detected in this study. Further studies of clefting modifiers and replication in larger, more diverse cohorts is warranted.

Although we initially focused on rare, coding variants we also found evidence that non-coding variants are important in modifying OFC subtypes. The gene bodies of *SEC24D* and *SUPT4H1* contained predicted craniofacial enhancers. The rare variants within these enhancers are predicted to alter TFBSs for many different TFs. Some, like Pax6 and PAX9, have been associated with orofacial clefts before (Peters et al. 1998; Rodrigo et al. 2003; Sull et al. 2009; Vaivads et al. 2021), and others, like PAX1, have been associated with both normal facial morphology and as laterality modifiers of orofacial clefts (Curtis et al. 2021a; Shaffer et al. 2016). This is consistent with the hypothesis that genetic variation in regulatory regions may have an effect by altering TFBSs and thus gene expression. However, for both genes in this study, the association with the coding variation within the gene was stronger than the association with all rare variants within the overlapping enhancer. Of the genes within their respective TADs, *SEC24D* and *SUPT4H1* were among the highest expressed genes during craniofacial development, making them likely target genes of these potential regulatory regions. Thus, while there is evidence consistent with rare, regulatory variants impacting orofacial cleft phenotypes, this effect is likely in addition to the effect by coding variation, and the effect of rare, non-coding variation genome-wide needs further investigation.

In summary, we conducted the first genome-wide scan of rare genetic modifiers in a case-case design for CL vs CLP and found that *SEC24D* and *SUPTAH1* had a significant burden of rare, coding variants in CL cases compared to CLP cases. By using publicly available datasets, we were able to determine that both genes were highly expressed in developing craniofacial tissue and contained predicted craniofacial enhancers. Additionally, we found that variants in these enhancers created or destroyed predicted binding sites for transcription factors, including *Pax9* and *Pax6*, suggesting a mechanism through which coding and noncoding variants could lead to phenotypic heterogeneity. Overall, this study expands our understanding of the genetic architecture of CL vs CLP and suggests that regulatory variation may also contribute and warrants further investigation in future studies.

## Data Availability

All data is available in the database of Genotypes and Phenotypes (dbGaP; European trios, dbGaP: phs001168.v2.p2; Colombian trios, dbGaP: phs001420.v1.p1; Taiwanese trios, dbGaP: phs000094.v1.p1) and from the Kids First Data Resource Center.

## Acknowledgments

The authors thank the dedicated field staff, collaborators, and participating families for their important contributions to this study. These studies are part of the Gabriella Miller Kids First Pediatric Research Program, supported by the Common Fund of the Office of the Director of the National Institutes of Health (NIH). Sequencing of the European trios was completed at Washington University’s McDonnell Genome Institute (3U54HG003079-12S1 and X01-HL132363 and the Colombian and Taiwanese trios were sequenced at the Broad Institute Sequencing Center (U24-HD090743). The sequencing centers plus the Kids First Data Resource Center, supported by the NIH Common Fund through grant U2CHL138346, provided technical and analytical support of this project. This work was supported by grants from the National Institutes of Health (NIH) including: F32-DE032260 [SWC], R00-DE025060 [EJL], R01-DE028342 [EJL], R03-DE030118 [EJL], R21-DE029698 [MPE], X01-HG007485 [MLM, EF], R01-DE016148 [MLM, SMW], U01-DE024425 [MLM], R37-DE008559 [JCM, MLM], R01-DE009886 [MLM], R21-DE016930 [MLM], R01-DE012472 [MLM], R01-DE014581 [THB], U01-DE018993 [THB], U01-DE020078; R01-DE027023 [SMW].

## Declarations

The authors have no disclosures.

## Data Availability

Phenotypes and associated genome sequencing data used in this manuscript are available from the database of Genotypes and Phenotypes (dbGaP; European trios, dbGaP: phs001168.v2.p2; Colombian trios, dbGaP: phs001420.v1.p1; Taiwanese trios, dbGaP: phs000094.v1.p1) and from the Kids First Data Resource Center.

