## Supplemental Figure for "Rare genetic variants in *SEC24D* modify orofacial cleft phenotypes"

**Supplementary Tables:**

Table S1: Cohort description

|  | RV-TDT analysis (CL) | RV-TDT analysis (CLP) | Modifier analysis |
| --- | --- | --- | --- |
| Number of samples | 228 | 675 | 369 |
| Number of families | 76 | 225 | 369 |
| Number of CL cases | 80 | 0 | 100 |
| Number of CLP cases | 0 | 241 | 269 |
| Number of males | 121 | 381 | 242 |
| Number of females | 107 | 294 | 127 |

Table S2: RV-TDT results for CL only (MAF < 1%; coding)

Table S3: RV-TDT results for CLP only (MAF < 1%; coding)

Table S4: Modifier results for CL vs CLP (MAF < 1%; coding)

Table S5: Annotation of variants in *SEC24D*

Table S6: Annotation of variants in *SUPT4H1*

Table S7: JASPAR results for *SEC24D* variants within enhancer regions

**Supplementary Figures:**

Figure S1: Q-Q plots of the −log_10_(p-values) of gene-based burden tests in the trios with CL only (A) and the trios with CLP only (B).

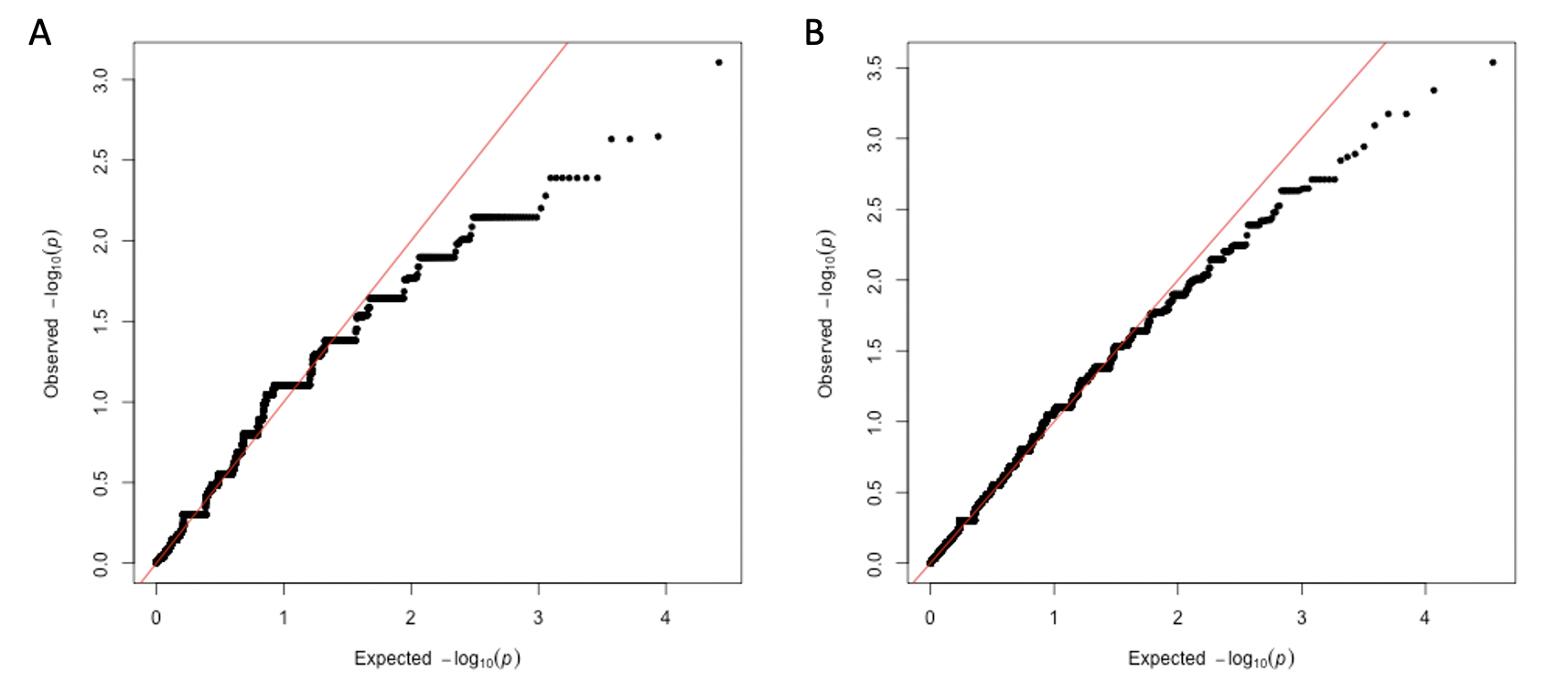

Figure S2: Manhattan plot of –log_10_(p-values) from the gene-based burden tests in the trios with CL only (A) and the trios with CLP only (B).

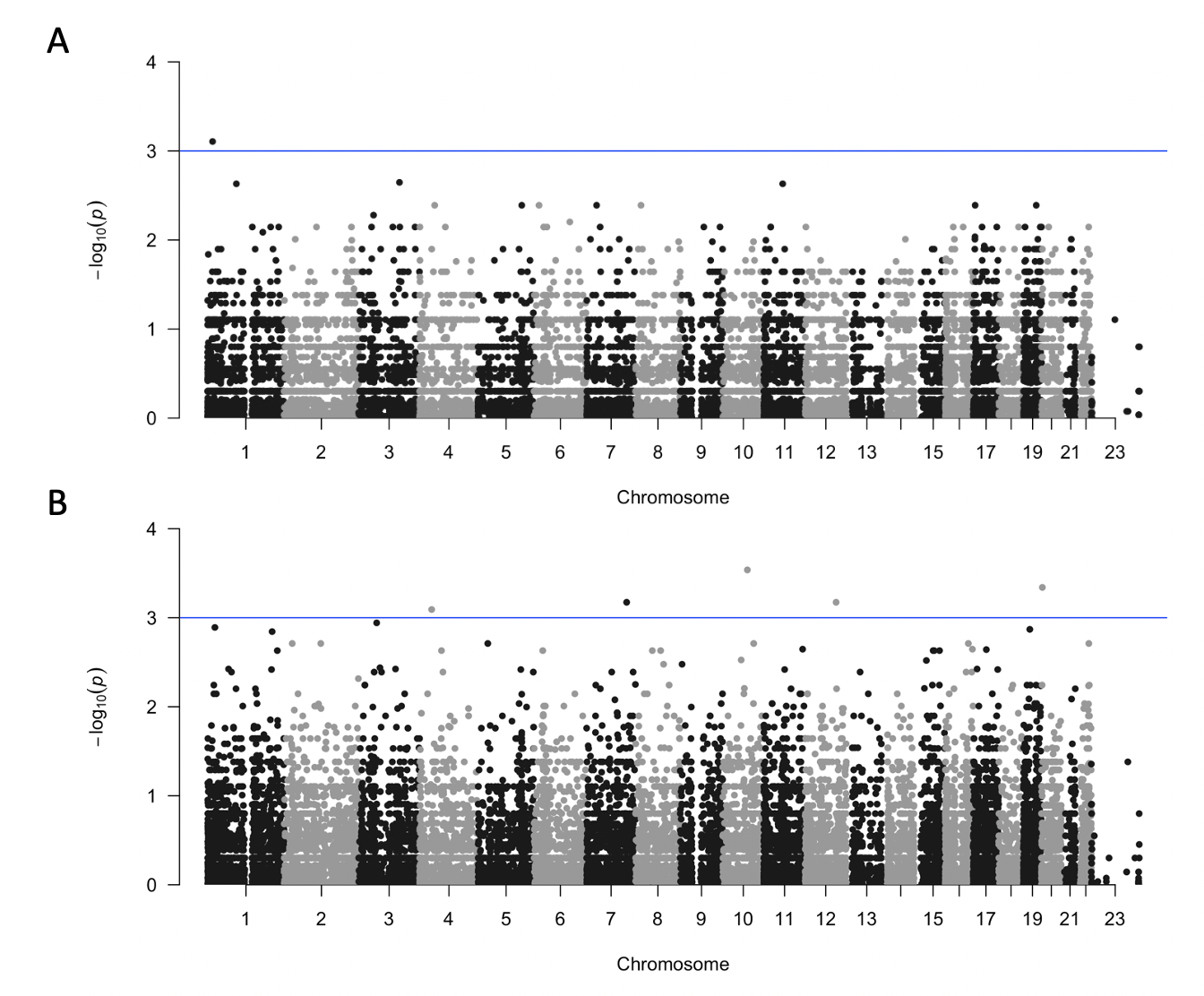

Figure S3: Comparison of the -log_10_(p-values) from the analysis with trios with CL only vs those from the analysis with trios with CLP only. There was no correlation between the two analyses (r = 0.005; p = 0.56)

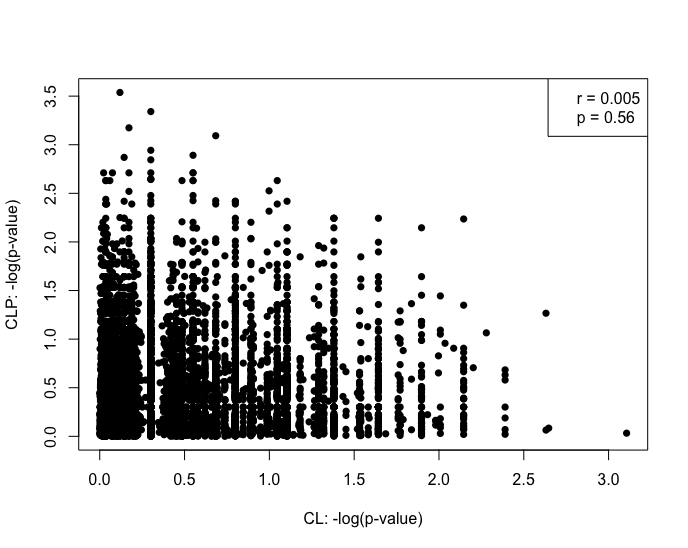

Figure S4: Q-Q plots of the −log_10_(p-values) of gene-based burden tests in the CL vs CLP modifier analysis. The genomic inflation factor is 1.04.

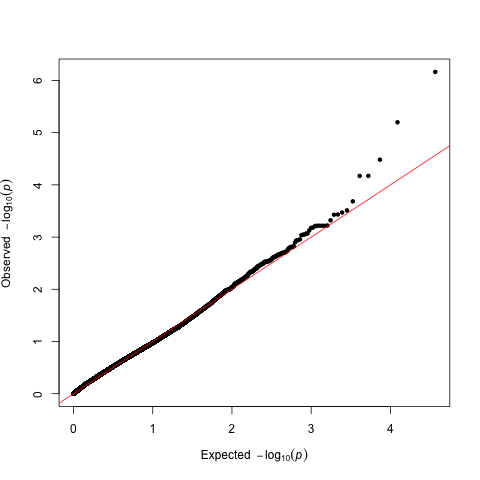

Figure S5: Rare variants within *SUPT4H1* fall within a predicted craniofacial super enhancer.

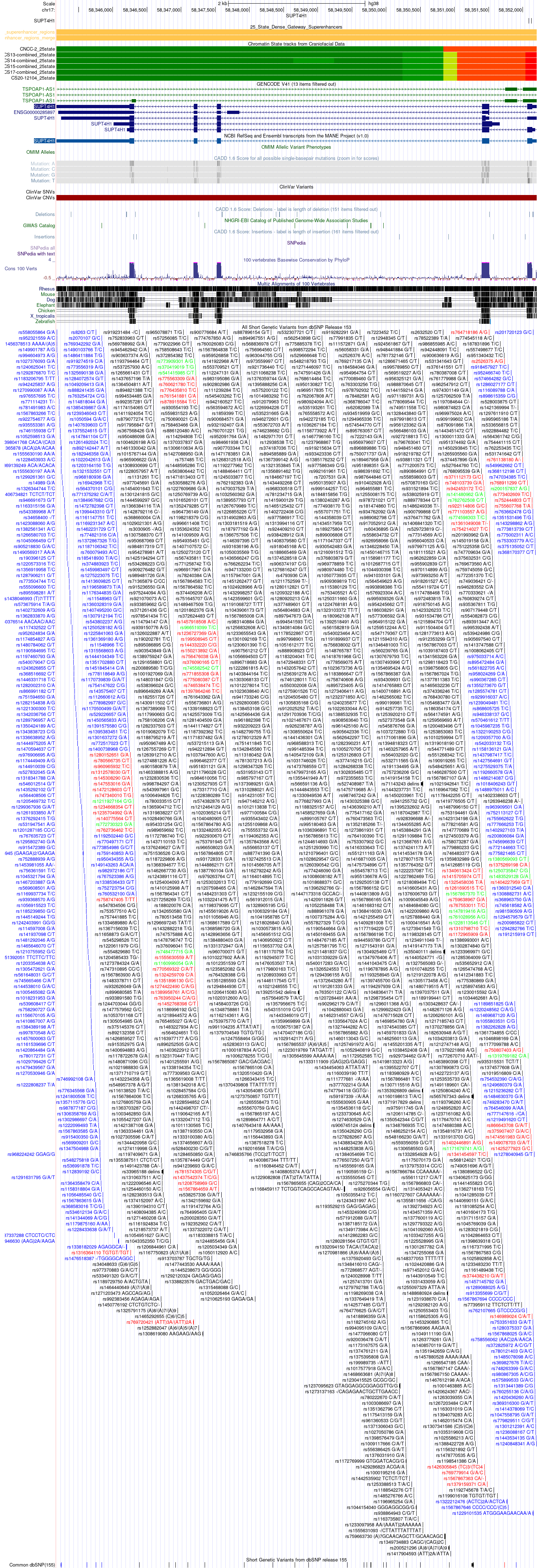

Figure S6: The predicted topologically-associated domain (TAD) surrounding *SUPT4H1*.
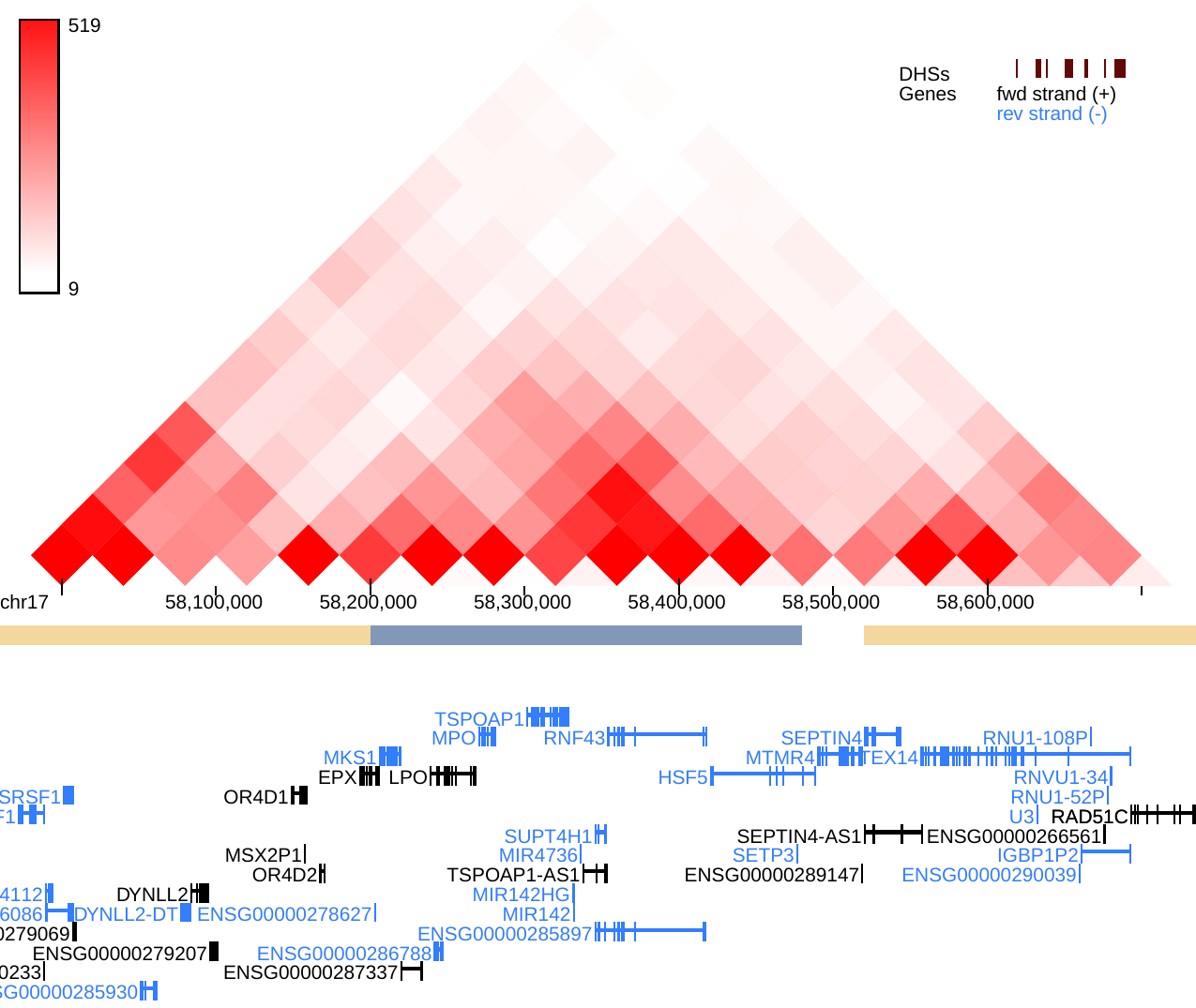
